# ‘Dark matter’, second waves and epidemiological modelling

**DOI:** 10.1101/2020.09.01.20185876

**Authors:** Karl J. Friston, Anthony Costello, Deenan Pillay

**Affiliations:** The Wellcome Centre for Human Neuroimaging, University College London; UCL Institute for Global Health, University College London; UCL Division of Infection and Immunity, University College London

## Abstract

**Background:** Recent reports based on conventional SEIR models suggest that the next wave of the COVID-19 pandemic in the UK could overwhelm health services, with fatalities that far exceed the first wave. These models suggest non-pharmaceutical interventions would have limited impact without intermittent national lockdowns and consequent economic and health impacts. We used Bayesian model comparison to revisit these conclusions, when allowing for heterogeneity of exposure, susceptibility, and viral transmission.

**Methods:** We used dynamic causal modelling to estimate the parameters of epidemiological models and, crucially, the evidence for alternative models of the same data. We compared SEIR models of immune status that were equipped with latent factors generating data; namely, location, symptom, and testing status. We analysed daily cases and deaths from the US, UK, Brazil, Italy, France, Spain, Mexico, Belgium, Germany, and Canada over the period 25-Jan-20 to 15-Jun-20. These data were used to estimate the composition of each country’s population in terms of the proportions of people (i) not exposed to the virus, (ii) not susceptible to infection when exposed, and (iii) not infectious when susceptible to infection.

**Findings:** Bayesian model comparison found overwhelming evidence for heterogeneity of exposure, susceptibility, and transmission. Furthermore, both lockdown and the build-up of population immunity contributed to viral transmission in all but one country. Small variations in heterogeneity were sufficient to explain the large differences in mortality rates across countries. The best model of UK data predicts a second surge of fatalities will be much less than the first peak (31 *vs*. 998 deaths per day. 95% CI: 24-37)—substantially less than conventional model predictions. The size of the second wave depends sensitively upon the loss of immunity and the efficacy of find-test-trace-isolate-support (FTTIS) programmes.

**Interpretation:** A dynamic causal model that incorporates heterogeneity of exposure, susceptibility and transmission suggests that the next wave of the SARS-CoV-2 pandemic will be much smaller than conventional models predict, with less economic and health disruption. This heterogeneity means that seroprevalence underestimates effective herd immunity and, crucially, the potential of public health programmes.

**Research in context:** *Evidence before this study:* Hundreds of modelling papers have been published recently, offering predictions and projections of the current coronavirus outbreak. These range from peer-reviewed publications to rapid reports from learned societies. Many, if not most, of these modelling initiatives commit to a particular kind of epidemiological model that precludes heterogeneity in viral exposure, susceptibility, and transmission. The ensuing projections can be fantastical in terms of fatalities and ensuing public health responses.

*Added value of this study:* This study revisits the evidence for conventional epidemiological modelling assumptions using dynamic causal modelling and Bayesian model comparison. It provides overwhelming evidence for heterogeneity, and the interaction between lockdown and herd immunity in suppressing viral transmission.

*Implications of all the available evidence:* Heterogeneity of this sort means that low seroprevalence (<20%) is consistent with levels of population immunity that play a substantive role in attenuating viral transmission and, crucially, facilitating public health measures.

## Introduction

The UK has suffered one of the highest death rates in the world from the second severe acute respiratory syndrome coronavirus (SARS-CoV-2). Three recent models project an even larger second wave of infections—with the UK facing an overwhelmed health service and death rates far higher than the first wave, unless a series of national lockdowns are enforced*^1,2^*. Okell and colleagues (*ibid*) suggest that ‘the epidemic is still at a relatively early stage and that a large proportion of the population therefore remain susceptible’. The Academy of Medical Science in their 14-July-20 report ^i^ ‘Preparing for a challenging winter 2020/21’ suggest a peak in hospital admissions and deaths in January/February 2021 with estimates of 119,900 (95% credible interval: 24,500-251,000) hospital deaths between September 2020 and June 2021—double the number that occurred during the first wave in the spring of 2020. Davies and colleagues^1^ project a median unmitigated burden of 23 million (95% CI: 13–30 million) clinical cases and 350,000 deaths (95% CI: 170,000–480,000) due to COVID-19 in the UK by December, 2021, with only national lockdowns capable of bringing the reproductive ratio near or below one. These kinds of projections have profound consequences for the national economy and the resulting health impacts of recession and unemployment.

This article challenges these projections and, in particular, the underlying assumptions that the risk of infection is homogeneous within the population. The role of pre-existing immunity, host genetics and overdispersion in nuancing viral transmission—and explaining the course of the pandemic in light of unlocking—calls for a more careful quantitative analysis^3-6^. The role of heterogeneity in exposure, susceptibility and transmission is receiving more attention; especially, in relation to the build-up of herd immunity^7,8^. This article illustrates a formal approach to epidemiological modelling that may help resolve some prescient issues.

The pessimistic projections above consider two principal mechanisms that underlie the mitigation—and possible suppression—of the ongoing coronavirus epidemic: (i) a reduction in viral transmission due to *lockdown* and social distancing measures, and (ii) a build-up of population or *herd immunity*. Herd immunity can be read as the population immunity that is required to attenuate community transmission. For example, Okell and colleagues (*ibid*) review three lines of argument and conclude that herd immunity is unlikely to explain differences in mortality rates across countries; thereby placing a strategic emphasis on lockdown to preclude a rebound of infections. This is in contrast with a herd immunity scenario, whereby immunity in the population will reduce transmission to pre-empt a second wave ^5,9,10^.

We use their analyses as a vehicle to question the validity of projections based upon conventional (susceptible, exposed, infected, and removed—SEIR) modelling assumptions. In particular, we deconstruct their arguments to show that the empirical observations they draw upon are consistent with herd immunity. Furthermore, public health responses and herd immunity are not mutually exclusive explanations for mortality rates, they both contribute to the epidemiological process and contextualise each other in potentially important ways. In turn, this has implications for the timing of interventions such as lockdown and FTTIS. More generally, we question the commitment to conventional epidemiological models that have not been subject to proper model comparison.

## Methods

Dynamic causal modelling is the application of variational Bayes to estimate the parameters of state-space models and, crucially, the evidence for alternative models of the same data. It was developed to model interactions among neuronal populations and has been used subsequently in radar, medical nosology and recently epidemiology^11-15^. Variational Bayes is also known as *approximate Bayesian inference* and is computationally more efficient than Bayesian techniques based upon sampling procedures (e.g., *approximate Bayesian computation*), which predominate in epidemiological modelling^16-18^. The particular dynamic causal model used here embeds an SEIR model of immune status into a model that includes all latent factors generating data; namely, location, infection symptom, and testing status. Please see the foundational paper for structural details of the model used in this paper^15^ and the generic (Variational Laplace) scheme used to estimate model parameters and evidence.

Dynamic causal modelling (DCM) differs from conventional epidemiological modelling in that it uses mean field approximations and standard variational procedures to model the evolution of probability densities^16^. This contrasts with epidemiological modelling that generally uses stochastic realisations of epidemiological dynamics to approximate probability densities with sample densities^17,19-21^. One advantage of variational procedures is that they are orders of magnitude more efficient; enabling end-to-end model inversion or fitting within minutes (on a laptop) as opposed to hours or days (on a supercomputer)^17^. More importantly, variational procedures provide an efficient way of assessing the quality of one model relative to another, in terms of model evidence (a.k.a., marginal likelihood) ^22^. This enables one to compare different models using Bayesian model comparison (a.k.a. structure learning) and use the best model for nowcasting, forecasting or, indeed, test competing hypotheses about viral transmission.

More generally, Bayesian model comparison plays a central role in testing hypotheses given (often sparse or noisy) data. It eschews intuitive assumptions, about whether there are sufficient data to test this or that, by evaluating the evidence for competing hypotheses or models. If there is sufficient information in the data to disambiguate between two hypotheses, the difference in log-evidence will enable one to confidently assert one model is more likely than the other. Note that this automatically ensures that the model is identifiable, in relation to the model parameters or prior assumptions in question.

Dynamic causal models can be extended to generate any kind of epidemiological data at hand: for example, the number of positive antigen tests. This requires careful consideration of how positive tests are generated, by modelling latent variables such as the bias towards testing people with or without infection or, indeed, the time-dependent capacity for testing. In short, everything that matters—in terms of the latent (hidden) causes of the data—can be installed in the model, including lockdown, self-isolation and other processes that underwrite viral transmission. Model comparison can then be used to assess whether the effect of a latent cause is needed to explain the data—by withdrawing the effect and seeing if model evidence increases or decreases. Here, we leverage the efficiency of dynamic causal modelling to evaluate the evidence for a series of models that are distinguished by heterogeneity or variability in the way that populations respond to an epidemic. The dynamic causal model used for the analyses below is summarised in terms of its structure (Figure 1) and parameters (Table 1):

**Figure 1:**
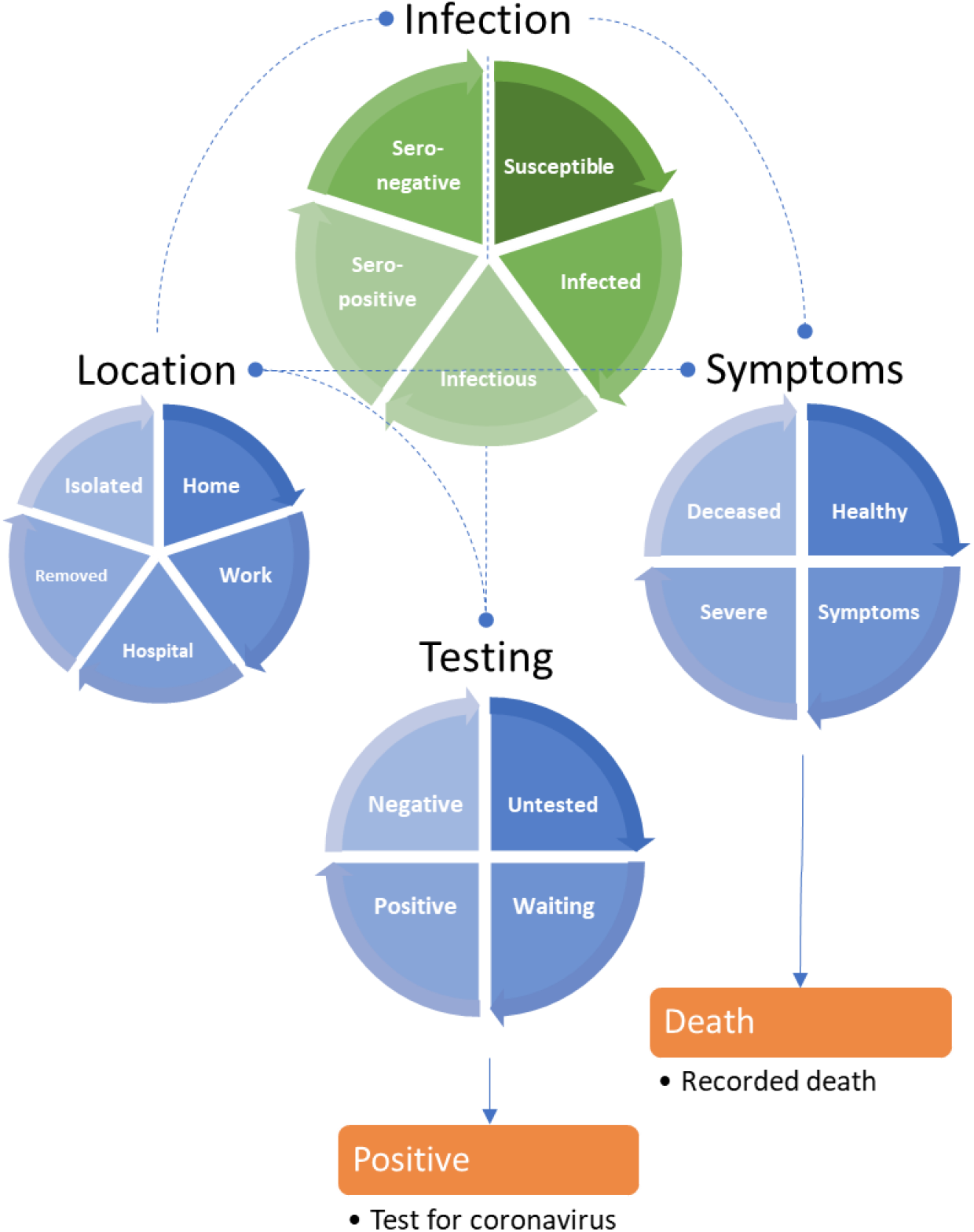
A LIST model. This schematic summarises a LIST (*location*, *infection, symptom*, and *testing*) model used for the accompanying analyses. This model is formally equivalent to the model in^23^. It includes a state (*isolated*) to model people who have yet to be exposed to the virus or are shielding because they think they may be infectious (within their home or elsewhere). It also includes a (*seronegative*) state to model individuals who have pre-existing immunity, e.g., via cross-reactivity ^24,25^ or other protective host factors ^26,27^. This absorbing state plays the role of the *recovered* or *removed* states of SEIR models—once entered, people stay in this state for the duration of the outbreak. One can leave any of the remaining states. For example, one occupies the *deceased* state for a day and then moves to *healthy* on the following day. Similarly, one occupies the state of testing *positive* or *negative* for a day, and then moves to the *untested* state the following day. This enables the occupancy of various states to be quantified in terms of daily rates. The discs represent the four factors of the model, and the segments correspond to their states (i.e., compartments). The green disc is the closest to a conventional (i.e., SEIR) model that is embedded within three other factors. The states within any factor are mutually exclusive. In other words, every individual has to be in one state associated with four factors. The orange boxes represent the observable outputs that are generated by this model, in this instance, daily reports of positive tests and deaths. The rate of transition between states—or the dwell time within any state—rests upon the model parameters that, in some instances, can be specified with fairly precise prior densities—listed in Table 1.

**Table 1.**
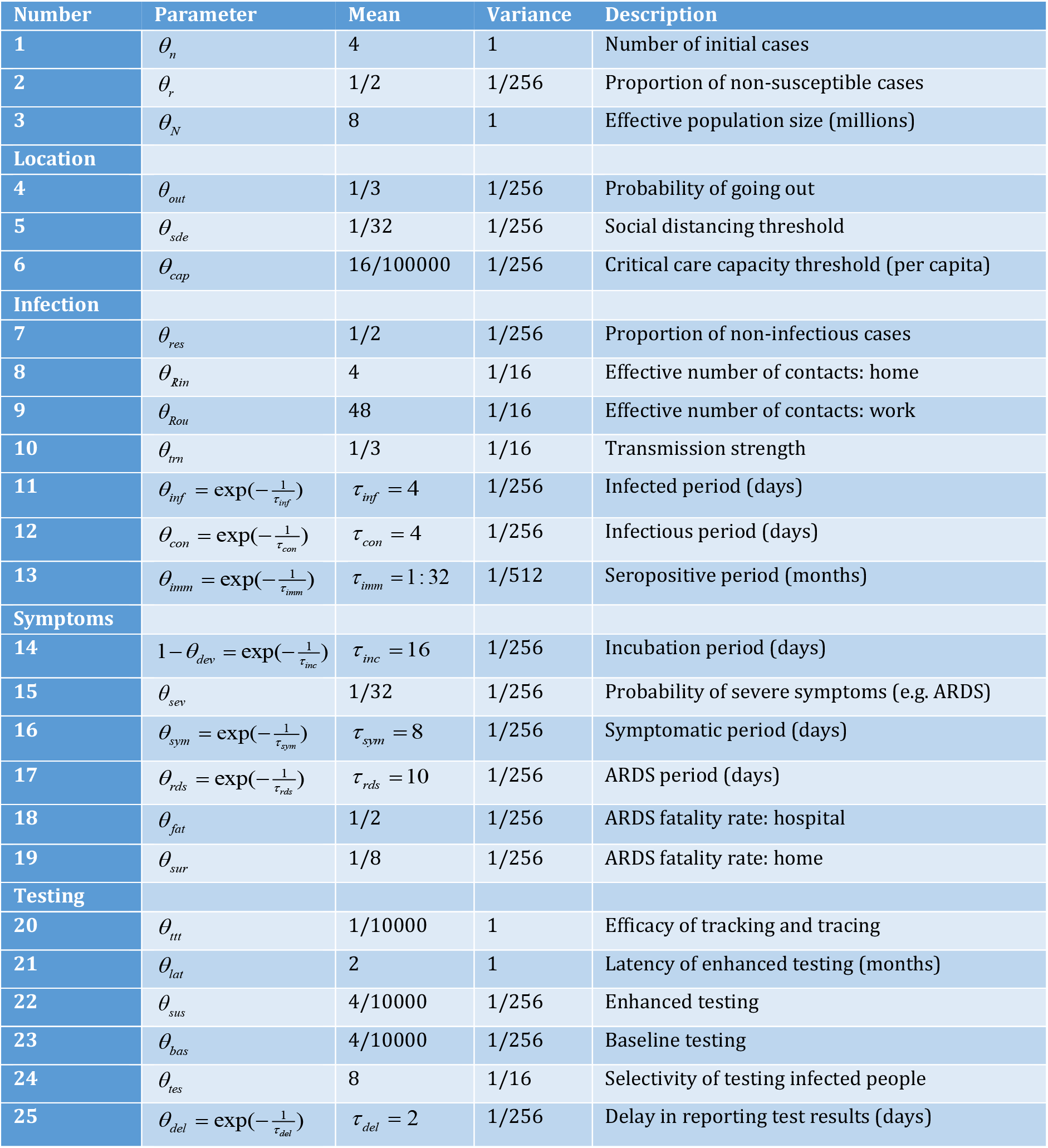
*Parameters of the epidemic (LIST) model and priors: N*(*η, C*) *(NB: prior means are for scale parameters θ =* exp(*ϑ*))

**Secondary sources** ^28-33^. The prior expectations should be read as the effective rates and time constants as they manifest in a real-world setting. The incubation period refers to the time constant corresponding to the rate at which one becomes symptomatic if infected—it does not refer to the period one is infected prior to developing symptoms. For example, early evidence indicates that by 14 days, approximately 95% of presymptomatic periods will be over ^34,35^. The priors for the non-susceptible and non-infectious proportion of the population are based upon clinical and serological studies reported over the past few weeks^36,37^. Please see the code base for a detailed explanation of the role of these parameters in transition probabilities among states. Although the (scale) parameters are implemented as probabilities or rates, they are estimated as log scale parameters, denoted by *ϑ* = ln *θ*. Note that the priors are over log scale parameters and are therefore mildly informative. For example, a prior variance of 1/256 corresponds to a prior standard deviation of 1/16. This means that the parameter in question can, *a priori*, vary by a factor of about 30%. The specific priors used for the current analyses are listed in **spm_COVID_priors.m**^a^ and can be optimised using Bayesian model comparison (by comparing the evidence with models that have greater or lesser shrinkage priors). Please see ^15^ for details.

Notice that this model is more nuanced than most conventional epidemiological models. For example, immunity and testing are separate factors. This means that we have not simply added an observation model to an SEIR-like model; rather, testing now becomes a latent factor that can influence other factors (e.g., the location factor via social distancing). Furthermore, there is a difference between the latent testing state and the reported number of new cases—that depends upon sensitivity and specificity, via thresholds used for reporting ^38^. Separating the infection and symptom factors allows the model to accommodate asymptomatic infection^39^: to move from an asymptomatic to asymptomatic state depends upon whether one is infected but moving from an infected to an infectious state does not depend upon whether one is symptomatic. Furthermore, it allows for viral transmission prior to symptom onset^40,41^.

This particular dynamic causal model accommodates heterogeneity at three levels that can have a substantive effect on epidemiological trajectories. These effects are variously described in terms of overdispersion, super-spreading, and amplification events^4,6,42^. In the current model, heterogeneity was modelled in terms of three bipartitions (summarised in Figure 2):

**Figure 2:**
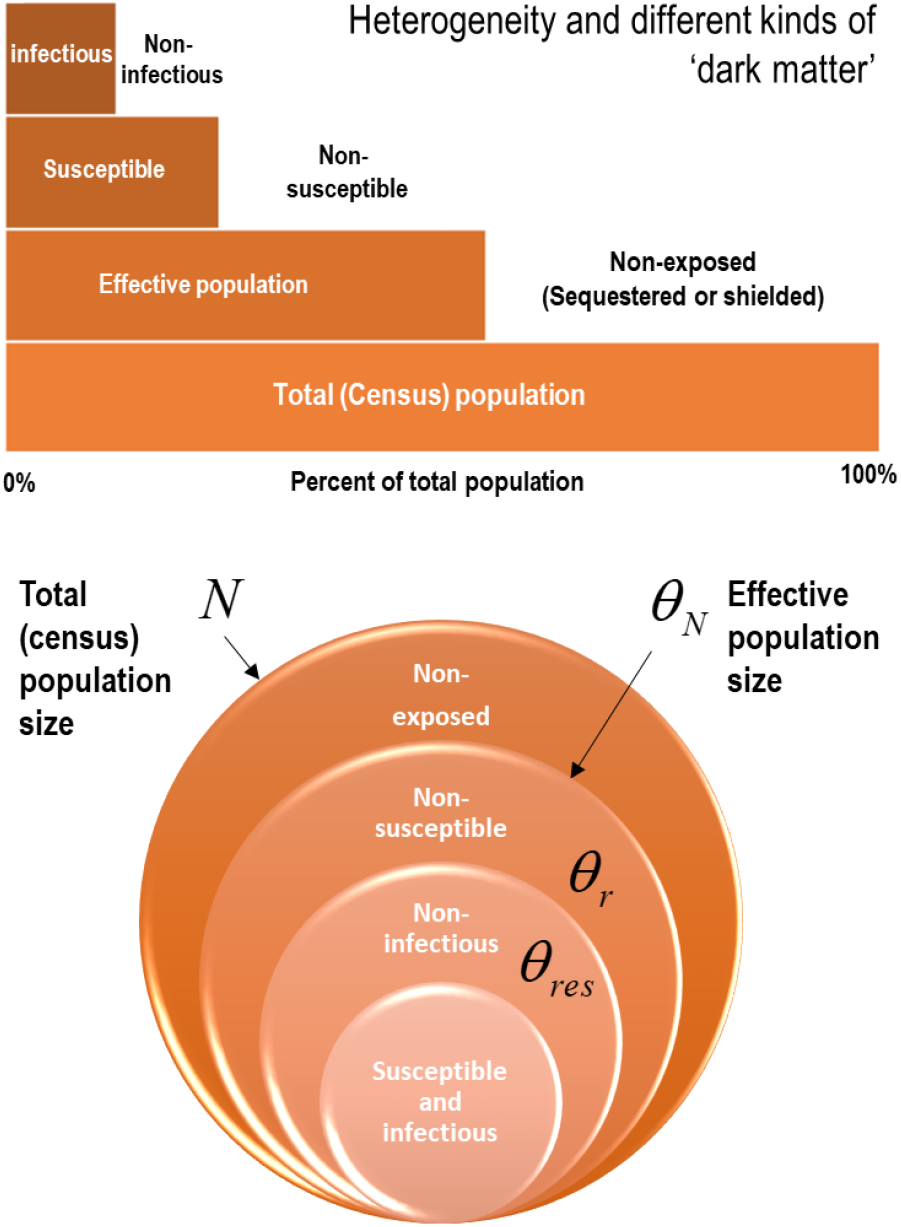
Heterogeneity of exposure, susceptibility, and transmission. U**pper panel**: this schematic illustrates the composition of a population in terms of a proportion that is not exposed to the virus, a proportion that is not susceptible when exposed and a further proportion of susceptible people who cannot transmit the virus. These proportions are unknown but can be estimated from the data. **Lower panel**: this schematic illustrates the parameterisation of heterogeneity in terms of the parameters in Table 1.

### Heterogeneity in exposure

This was modelled in terms of an effective population size that is less than the total (census) population. The effective population comprises individuals who are in contact with other infected individuals. The remainder of the population are assumed to be geographically sequestered from a regional outbreak or are shielded from it. For example, if the population of the UK was 68 million, and the effective population was 39 million, then only 57% are considered to participate in the outbreak. Of this effective population, a certain proportion are susceptible to infection:

### Heterogeneity in susceptibility

This was modelled in terms of a portion of the effective population that are not susceptible to infection. For example, they may have pre-existing immunity via cross- reactivity^24,25,43^ or particular host factors^26,27^ such as mucosal immunity^44^. This non-susceptible proportion is assigned to the *seronegative* state at the start of the outbreak. Of the remaining susceptible people, a certain proportion can transmit the virus to others:

### Heterogeneity in transmission

We modelled heterogeneity in transmission with a free parameter (with a prior of one half and a prior standard deviation of 1/16). This parameter corresponds to the proportion of susceptible people who are unlikely to transmit the virus, i.e., individuals who move directly from a state of being *infected* to a *seronegative* state (as opposed to moving to a *seropositive* state after a period of being *infectious*). We associated this transition with a mild infection^45^ that does not entail seroconversion, e.g., recovery in terms of T-cell mediated responses^25,26^. In short, the *seronegative* state plays the role of a *seropositive* state of immunity for people who never become infectious, either because they are not susceptible to infection or have a mild infection (with or without symptoms, e.g., children).

Modelling heterogeneity of susceptibility in terms of susceptible and non-susceptible individuals can be read as modelling the difference between old (susceptible) and young (non-susceptible) people^29^. However, in contrast to models with age-stratification, the current model does not consider different contact rates between different strata (e.g., contact matrices). Instead, we model variations in contact with location—in terms of the number of people one is exposed to in different locations. Although full stratification is straightforward to implement^b^, this simplified model of heterogeneity is sufficient to make definitive inferences about the joint contribution of lockdown and population immunity to viral transmission.

This kind of model is sufficiently expressive to reconcile the apparent disparity between morbidity/mortality rates and low seroprevalence observed empirically ^36,ii^. We will see below that Bayesian model comparison suggests there is very strong evidence ^46^ for all three types of heterogeneity.

Given a suitable dynamic causal model one can use standard variational techniques to fit the empirical data and estimate model parameters. Having estimated the requisite model parameters, one can then reconstitute the most likely trajectories of latent states: namely, the probability of being in different locations, states of infection, symptom, and testing states. An example is provided in Figure 3 using daily cases and death-by-date data from the United Kingdom from 30-Jan-20 to 01-Sep-20.

**Figure 3:**
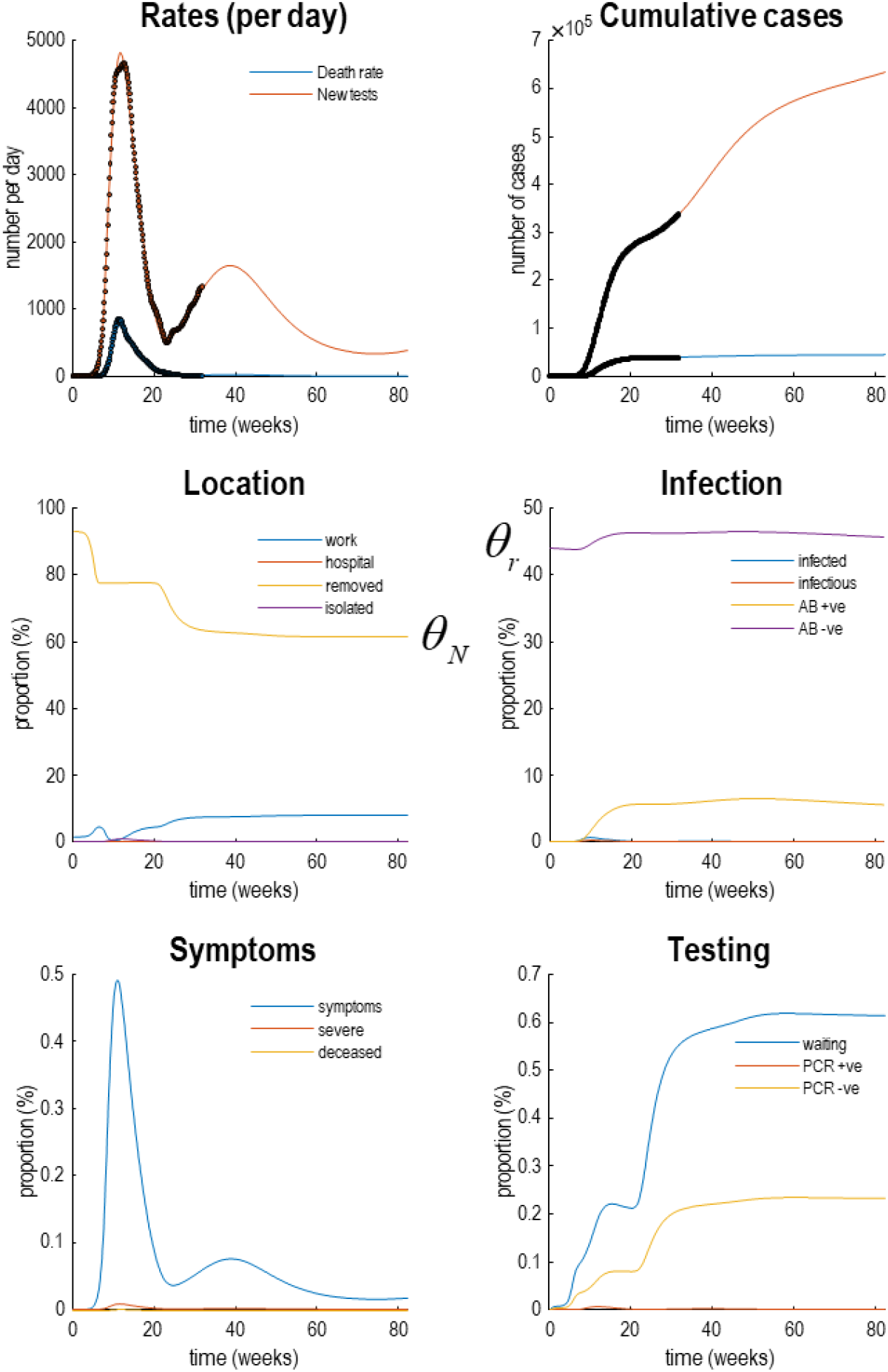
Latent causes. This figure illustrates posterior predictions of the most likely latent states for the United Kingdom. Here, the outcomes in the upper two panels (dots) are supplemented with the underlying latent causes or expected states in the lower four panels (the first state in each factor has been omitted for clarity: i.e., *home*, *susceptible*, *healthy*, and *untested*). These latent or expected states generate the observable outcomes in the upper two panels. The solid lines are colour-coded and correspond to the states of the four factors in **Error! Reference source not found**.. For example, under the *location* factor, the probability of being found at in a high-risk location (*work*: blue line) rises at the onset of the outbreak, falls during lockdown, and then slowly recovers during unlocking. At the same time, the proportion of the population who are not exposed to the virus (*removed*: yellow line) falls, as the exposed proportion increases to the effective population size. During lockdown, the probability of isolating oneself rises to about 3% during the peak of the pandemic (*isolated*: purple line). After about six weeks, lockdown starts to relax and slowly tails off, with accompanying falls in morbidity (in terms of *symptoms*: blue line in the *symptom* panel) and mortality (in terms of death rate). Note that seroprevalence (*Ab +ve*: yellow line in the *infection* panel) peaks at about 50 weeks and then starts to decline. Please see software and data note for details about the model inversion and data used to prepare this figure—and Figure 6 for confidence intervals on fatality rates.

## Results and commentary

Having briefly established the form and nature of the quantitative modelling, we now apply it to daily reports of new cases and deaths from several countries. Our focus is not on the details of the model—or its predictions. Rather, we use the modelling to illustrate how Bayesian model comparison can be used to test some assumptions that underwrite conventional models. In the setting, we frame the results in the form of a commentary and restrict the analysis to data available at the time the above reports were published (i.e., from 25-Jan-20 to 15-June-21).

### Heterogeneity in exposure, susceptibility, and transmission

In what follows, we use dynamic causal modelling to revisit some assumptions implicit in conventional epidemiological modelling. We follow the three lines of arguments rehearsed in Okell et al (ibid). The first can be summarised as: *under herd immunity the cumulative mortality rate per million of the population should plateau at roughly the same level in different countries*.

This is true if, and only if, the same proportion of the population can transmit the virus. In other words, a plateau to endemic equilibrium—based on the removal of susceptible people from the population—only requires people are susceptible to infection to be immune. If this proportion depends upon the composition of a country’s population (i.e., demography), mortality rates could differ from country to country. This can be illustrated by using models with heterogeneous population structures, of the sort summarised in Figure 2. Figure 4 shows the data and ensuing predictions for ten countries, using the format of Okell et al.^2^

**Figure 4:**
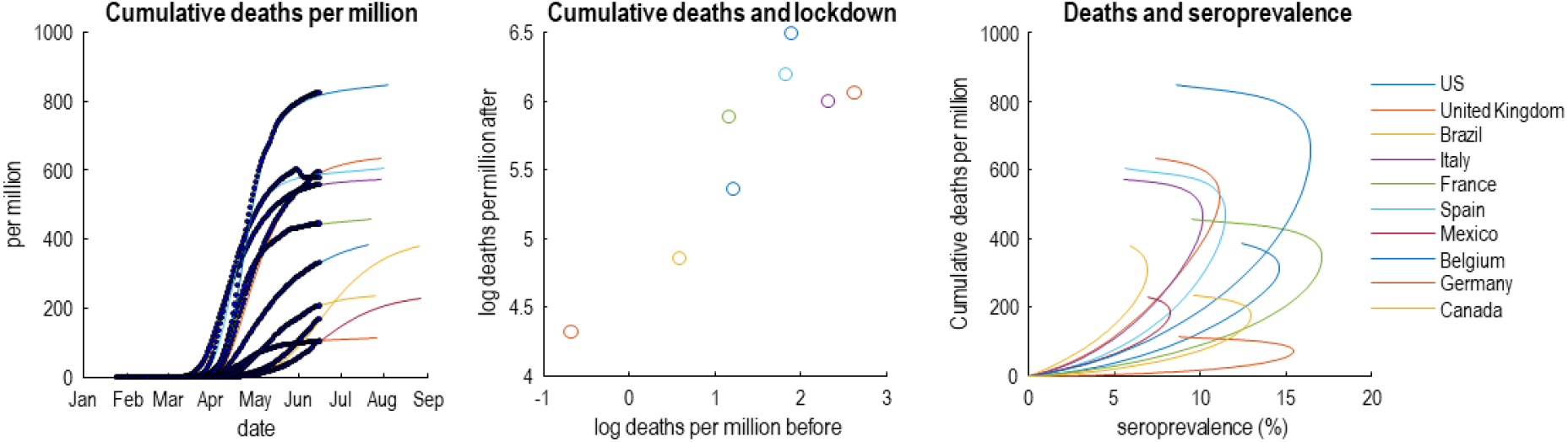
This figure reproduces the quantitative results in the Okell et al (ibid). However, here, they are based upon the predictions of a dynamic causal model of the epidemic in the 10 countries with the highest mortality on 15-Jun-20. The left panel shows the cumulative deaths per million as a function of time to illustrate variation over countries. The middle panel plots the log deaths per million in the six weeks following lockdown against the corresponding mortality before lockdown. The right panel plots the estimated cumulative deaths against estimated seroprevalence in the effective population over a period of 180 days. Note that these graphics include both empirical data (dots and circles) and predictions (coloured lines) based upon the parameter estimates of the dynamic causal modelling (credible intervals have been omitted for clarity). The predictions of seroprevalence are, in this example, based purely upon new cases and deaths. Although it would be relatively straightforward to include empirical seroprevalence data in the dynamic causal modelling, we have presented the seroprevalence predictions to illustrate the predictive validity of the model—given that the predictions are in line with empirical reports (i.e., between 5% and 20%).

Here, we modelled new cases and COVID-19 related deaths in several countries with an effective population of unknown size. The remaining, non-exposed, proportion of the total (census) population are taken to be geographically sequestered or shielded from exposure to infection. The effective population comprises a mixture of susceptible and non-susceptible individuals with pre-existing protection from prior exposure to non-COVID-19 coronaviruses or other host protective characteristics^24-27,43,44^. In reality, this bipartition stands in for a graded susceptibility within the population. In turn, susceptible individuals comprise a mixture of people who are able and unable to transmit virus following infection (see Figure 2). The latter proportion may have a short period of virus shedding and low viral load, rendering them, in effect, non-infectious^45^. These non-infectious individuals are assumed to have a mild infection that does not entail seroconversion, e.g., recovery in terms of T-cell mediated responses^25,26,47^. This kind of model accommodates heterogeneity of exposure, susceptibility, and transmission (portrayed as ‘immunological dark matter’ in the media^iii^). Crucially, slight changes in the composition of the ensuing population can produce substantial variations in fatality rates (in the left panel of Figure 4).

Heterogeneity of transmission may be particularly important here^3^. This is sometimes framed in terms of overdispersion or the notion of superspreading and amplification events^4,6,42^. For example, if only 20% of the population were able to develop a sufficient viral load to infect others, then protective immunity in this subpopulation would be sufficient for an innocuous endemic equilibrium^48^. Furthermore, if seroconversion occurs largely in the subpopulation spreading the virus^49^, a sufficient herd immunity may only require a seroprevalence of around 10% of the effective population (right panel of Figure 4). In short, one might challenge the assumption that COVID-19 is spread homogeneously across the population. Indeed, heterogeneity is becoming more and more evident in high risk settings, and in the variation in the period of infectivity across ages.

This begs the question: is there evidence for heterogeneity in the dispersion of SARS-CoV-2? And, if so, does this heterogeneity vary from country to country? Figure 5 answers this question using Bayesian model comparison. It shows—under the models in question—there is overwhelming evidence for heterogeneity of exposure, susceptibility, and transmission. And that a substantial proportion of each country’s population does not contribute to viral transmission. These proportions vary from country to country, leading to the differential mortality rates in Figure 4.

**Figure 5:**
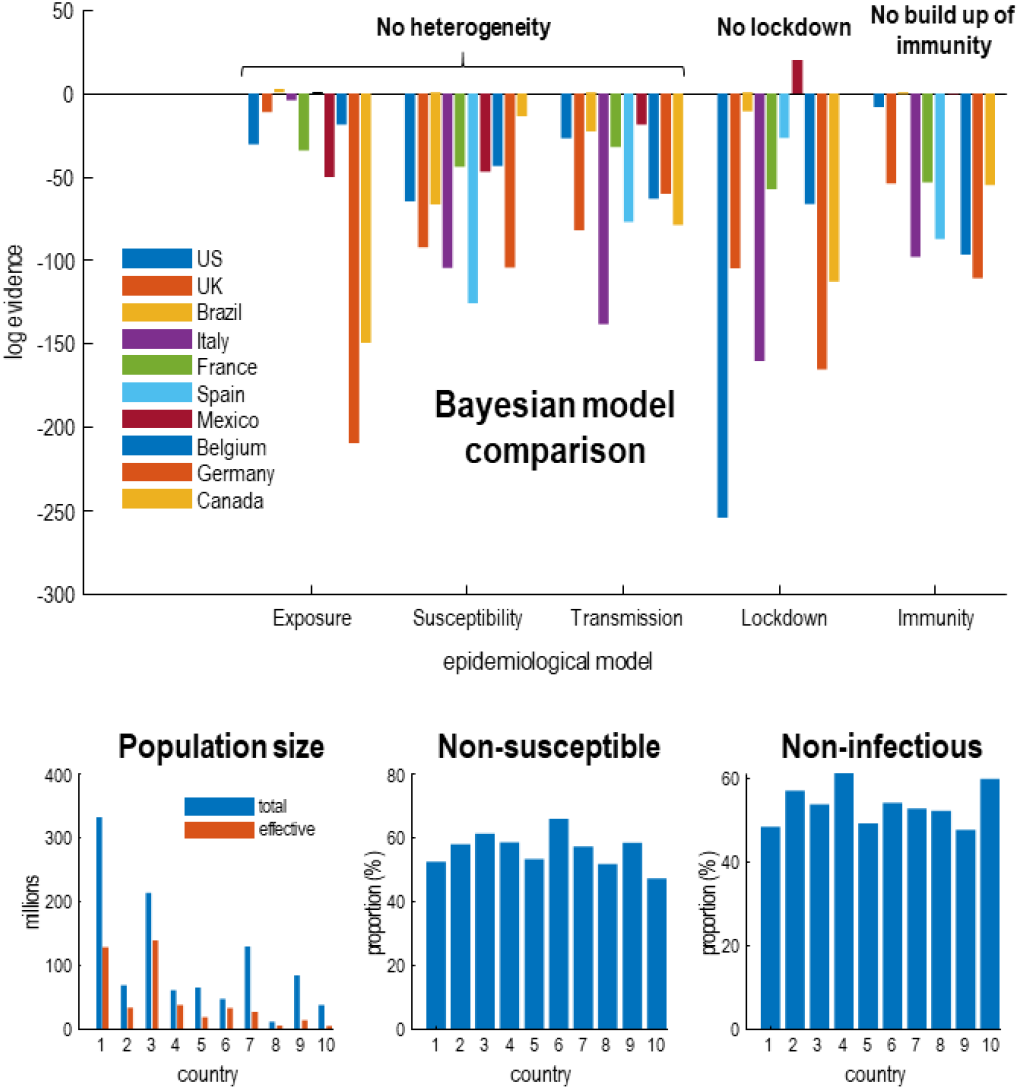
Bayesian model comparison of dynamic causal models of the coronavirus outbreak in 10 countries with the greatest mortality rates. The upper panel shows the drop in log evidence when removing a component from the model. This reduction is known as log Bayes factor. For example, if the **No build-up of herd immunity** model for the UK has a log evidence of -54, then it is *exp*(54) times less likely than the full model that includes a build-up of immunity. By convention, a difference in log evidence of 3 is taken to indicate strong evidence in favour of the full model. The full model accommodates heterogeneity at three levels. The first considers an effective population that is exposed to infection, where the remainder are geographically isolated or shielding. Of this effective population, a certain proportion cannot be infected (e.g., due to host factors such as pre-existing immunity). Of the remaining susceptible population, a certain proportion cannot transmit the virus (e.g., through having a short period of viral shedding). This leaves a subpopulation who are both susceptible to infection and capable of transmission. By removing these three kinds of heterogeneity—and re-evaluating the evidence for the reduced models in relation to the full model—one can assess the evidence for the contribution of heterogeneity. Similarly, one can remove social distancing and the build-up of seropositive immunity from the model and evaluate their respective contribution. The results in the **upper panel** indicate very strong evidence in favour of the full model for nearly all countries. The exceptions are Brazil and Spain—where the entire population appears to participate in the outbreak—and Mexico, where social distancing is not evident. Mexico is an interesting example where removing a model parameter increases model evidence (via a reduction model complexity). *Full model*: this model incorporates all three forms of heterogeneity, social distancing, and seropositive immunity. *Exposure*: heterogeneity to exposure was removed by equating the effective and total population. *Susceptibility*: heterogeneity of susceptibility was reduced by decreasing the prior proportion of non-susceptible people by a factor of *e*. *Transmission*: heterogeneity of transmission was reduced by decreasing the prior proportion of non-infectious people by a factor of *e*. *Lockdown*: social distancing was reduced by increasing the social distancing threshold by a factor of *e*^2^. *Immunity*: the effect of immunity was reduced by decreasing the period of seropositivity by a factor of *e*^2^. The **lower panels** provide the posterior expectations (i.e., most likely value given the data) of the effective population under the full model, alongside the census population (left panel). The subsequent panels show the non-susceptible proportion of the effective population (middle panel) and non-infectious proportion of the susceptible population (right panel). Credible intervals have been submitted for clarity but were in the order of 10 to 20%.

In general, the effective population is roughly half of the census population, with some variation over countries. The non-susceptible and non-infectious proportions are roughly half of the effective and susceptible populations, respectively—varying between 45% and 65%. This variation underwrites the differences in fatality rates in Figure 4. It could be argued that the estimates of the proportion of non-susceptible individuals is at odds with empirical data from contained outbreaks. For example, on the aircraft carrier Charles de Gaulle, about 70% of sailors were infected^c^. However, this argument overlooks the contribution of heterogeneity: there were no children on the Charles de Gaulle.

Okell et al (ibid) wrote: “If acquisition of herd immunity was responsible for the drop in incidence in all countries, then disease exposure, susceptibility, or severity would need to be extremely different between populations.” On the basis of the above quantitative modelling, this assumption transpires to be wrong: small variations in heterogeneity of exposure and susceptibility are sufficient to explain the differences between countries. Our point here, is that predicates or assumptions of this sort can be evaluated quantitatively in terms of model evidence.

### Do countries that went into lockdown early experience fewer deaths in subsequent weeks?

This is the second argument made by Okell et al (ibid) for the unique role of lockdown in mitigating fatalities: However, exactly the same correlation—between cumulative deaths before and after lockdown—emerges under epidemiological models that entertain heterogeneity and herd immunity. See Figure 4 (middle panel). In short, had the authors tested the hypothesis that lockdown or herd immunity were necessary to explain the data, they would have found very strong evidence for both—and may have concluded that lockdown nuances the emergence of herd immunity. See Figure 5.

### Does a correlation between antibodies to SARS-CoV-2 (i.e., seroprevalence) and COVID-19 mortality rates imply a similar infection fatality ratio (IFR) over countries?

Conventional models generally assume this is the case^2,29^. The problem with this assumption is that it precludes pre-existing immunity and loss of immunity (as in SEIRS models)^24^. Population immunity could fall over a few months due to population flux and host factors, such as loss of neutralising antibodies^49^. This is important because it means that seroprevalence could fall slowly after the first wave of infection (indeed, empirical seroprevalence is not increasing and may be decreasing in the UK ^iv^). In turn, this produces a nonlinear relationship between the prevalence of antibodies and cumulative deaths at the time seroprevalence is assessed. This is illustrated by the curvilinear relationships in the right panel of Figure 4 (under a loss of seroprevalence with a time constant of three months). If one associates IFR with the slope of fatality rates—as a function of seroprevalence—then the IFR changes over time. In short, the IFR changes as the epidemic progresses, as the proportion of susceptible and transmitting people falls, or those at highest risk succumb early in the pandemic. This begs the question: are quantities such as IFR fit for purpose when trying to model epidemiological dynamics?

### What is the impact of different rates of loss of immunity?

So, what are the implications of heterogeneity for seroprevalence and a second wave? The Bayesian model comparisons in Figure 5 speak to a mechanistic role for herd immunity in mitigating a rebound of infections. Note that this model predicts seroprevalences that are consistent with empirical community studies, *without ever seeing these serological data* (e.g., in the UK if 11% of the effective population is seropositive and the effective population is 49% of the census population, we would expect 5.4% of people to have antibodies, which was the case at the time of analysis^v^).

Predictive validity of this sort generally increases with model evidence. This follows from the fact that the log-evidence is accuracy minus complexity. In other words, models with the greatest evidence afford an accurate account of the data at hand, in the simplest way possible. Unlike the Akaike and widely used Bayesian information criteria (AIC and BIC), the variational bounds on log-evidence used in in dynamic causal modelling evaluate complexity explicitly^22^. Models with the greatest evidence have the greatest predictive validity because they do not overfit the data. An example of the posterior predictions afforded by the current model is provided in Figures 3 and 6 that speak to the timing and amplitude of a second wave in the UK. Figure 6 focuses on fatality rates under a couple of different scenarios; namely, under a rapid loss of antibody-mediated immunity and under and accelerated FTTIS program.

**Figure 6:**
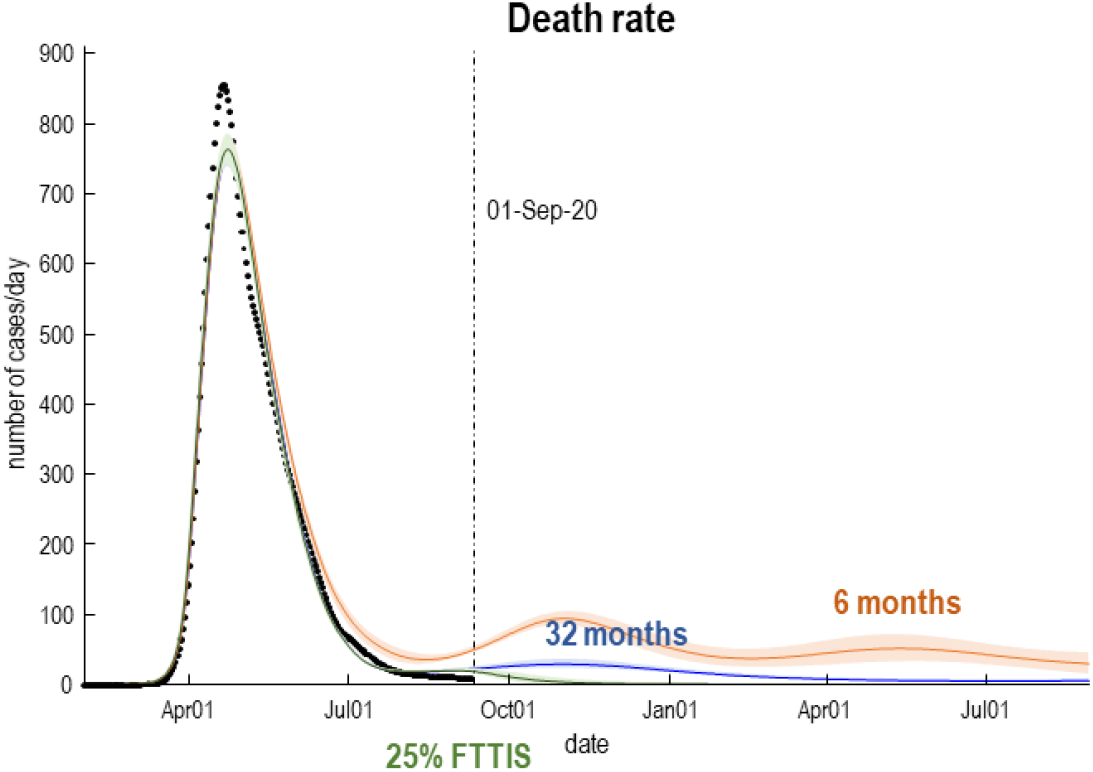
Expected death rates as a function of time for a heterogeneous United Kingdom population. The trajectories (lines) and accompanying 90% Bayesian credible intervals (shaded areas) correspond to posterior predictions with a loss of immunity over 32 (blue) and 6 (orange) months. This subsumes loss of protective immunity due to population fluxes and the immunological status of individuals. The six-month scenario can be considered a worst-case prediction, under which we would expect a second peak of about 100 deaths per day. Under the more optimistic prior of ensuring immunity fatality rates would be substantially lower than witnessed during the first peak (31 versus 856 per day, respectively. CI: 24-37). The green line reproduces the predictions based upon the posterior parameter but increasing the efficacy of find, test, trace, isolate and support to 25% (from its posterior estimate of about 1%).

These posterior predictions suggest that, under the assumption that immunity endures for a year or two, there may be a mild inflation of fatality rates over the autumn, peaking at about 30 per day. This second wave could be eliminated completely with an increase in the efficacy of contact tracing (FTTIS)—modelled as the probability of self-isolating, given one is infected but asymptomatic. It can be seen that even with a relatively low efficacy of 25%, elimination is possible by November, with convergence to zero fatality rates. Please see ^50^ for a more comprehensive analysis. Note that in a month or two, death rates should disambiguate between these scenarios.

The credible intervals in Figure 6 may appear rather tight. This reflects two issues. First, the well-known overconfidence problem with variational inference—discussed in ^15,51^. Second, these posterior predictive densities are based upon the entire timeseries, under a dynamic causal model that constrains the functional form of the trajectories. Put simply, this means that uncertainty about the future can be reduced substantially by data from the past.

## Discussion

Our reading of the epidemiological modelling literature suggests a systemic failure to formally evaluate the evidence for alternative models (e.g., models with age stratification and heterogeneous contact structure). This may reflect the fact that agent-based, stochastic transmission models are notoriously difficult to evaluate in terms of their evidence^16,18^. In contrast, the variational approaches used in dynamic causal modelling^34-39^ furnish a variational bound on model evidence that allows competing models to be assessed quickly and efficiently. The central role of model comparison is established in many disciplines and is currently attracting attention in epidemiology^52^.

Although model evaluation using the AIC (or widely used BIC) can be found in the epidemiological literature^53-55^, this kind of comparison does not constitute proper model comparison. This is because the complexity part of model evidence is not estimated by the AIC (or widely used BIC). The model complexity corresponds to the degrees of freedom used to explain the data (technically, the KL divergence between the posterior and prior). The AIC and BIC approximate complexity with (functions of) the number of free parameters, irrespective of whether these parameters are used or not. This means that the AIC is not fit for purpose when comparing models in a clinical or epidemiological setting. Please see ^22^ for illustrations of the failure of the AIC (and BIC). In short, it appears that most of the predictions underwriting ‘scientific advice’ to governmental agencies is based on epidemiological models that have not been properly compared with alternative models. If there is no rebound in fatality rates in the next few months, the conclusions in Okell et al, Davies et al and the Academy of Medical Sciences report (*ibid*) will be put under some pressure. This pressure might license a more (model) evidence-based approach, using the requisite variational methods that predominate in statistical physics, machine learning and (dynamic) causal modelling.

The recurrent theme above is the danger of committing to one particular model or conception of the epidemiological process. In other fields—dealing with population dynamics—Bayesian model comparison is used to identify the best structure and parameterisation of models^28-30^ known as structure learning^31,32^. Figure 5 offers an example of Bayesian model comparison in epidemiology, evincing very strong evidence for heterogeneity in responses to viral infection—and a synergistic role for social distancing and herd immunity.

Identifying the right epidemiological model has considerable public health and economic implications. While SARSCo-V2 may not be eradicated, model selection suggests that any second wave will be much smaller than other models have projected, and the virus will become endemic rather than epidemic. The size of a second wave may depend sensitively on the efficacy of FTTIS programmes and the rate of loss of immunity. Recent evidence suggests T-cell immunity may be more important for longer term immunity with circulating SARS-CoV-2-specific CD8+ and CD4+ T cells identified in 70% and 100% of COVID-19 convalescent patients, respectively^25^. Furthermore, 90% of people who seroconvert make detectible neutralizing antibody responses that are stable for at least 3 months^56^. If the above dynamic causal model is broadly correct, future national lockdowns may be unnecessary. As an endemic and potentially fatal virus, especially in elderly people and those with underlying conditions, attention to the details of FTTIS and shielding becomes all the more important. This emphasises the need for clear criteria for when and how to implement local lockdowns in ‘hotspot’ areas.

In summary, lockdown and social distancing have undoubtedly restricted the transmission of the virus. Model comparison suggests that these approaches remain an essential component of pandemic control, particularly at current levels of infections in the UK. However, extending the notion of ‘herd immunity’—to include seronegative individuals with lower susceptibility and/or lower risk of transmission—engenders an immune subpopulation that can change over time and country. The implicit immunity may reduce mortality and lower the risk of a second wave to a greater extent than predicted under many epidemiological models. On this view, herd immunity subsumes people who are not susceptible to infection or, if they are, are unlikely to be infectious or seroconvert; noting that SARS-CoV-2 can induce virus-specific T-cell responses without seroconversion. This reconciles the apparent disparity between reports of new cases, mortality rates and the low seroprevalence observed empirically. Crucially, Bayesian model comparison confirms that there is very strong evidence for the heterogeneity that underwrites this kind of herd immunity.

Put simply, an effective herd immunity—that works hand-in-hand appropriate public health and local lockdown measures—requires less than 20% seroprevalence. This seroprevalence has already been reached in many countries and is sufficient to preclude a traumatic second wave, even under pessimistic assumptions about loss of humoral immunity endowed by antibodies.

## Data Availability

The analyses in this article can be reproduced using annotated (MATLAB/Octave) code available as part of the free and open source academic software SPM (https://www.fil.ion.ucl.ac.uk/spm/), released under the terms of the GNU General Public License version 2 or later. The routines are called by a demonstration script: DEM_COVID_I.m. Please visit https://www.fil.ion.ucl.ac.uk/spm/covid-19/. The data used in this article are available for academic research purposes from the COVID-19 Data Repository by the Center for Systems Science and Engineering (CSSE) at Johns Hopkins University, hosted on GitHub at https://github.com/CSSEGISandData/COVID-19.

https://www.fil.ion.ucl.ac.uk/spm/covid-19/

https://github.com/CSSEGISandData/COVID-19/

## Glossary of terms

**Dynamic causal modelling**: the application of variational Bayes to estimate the unknown parameters of state-space models and assess the evidence for alternative models of the same data. See http://www.scholarpedia.org/article/Dynamic_causal_modeling.
**Variational Bayes** (a.k.a., **approximate Bayesian inference**): a generic Bayesian procedure for fitting and evaluating generative models of data by optimising a **variational bound** on **model evidence**. See https://en.wikipedia.org/wiki/Variational_Bayesian_methods.
**Model evidence** (a.k.a. **marginal likelihood**): the probability of observing some data under a particular model. It is called the marginal likelihood because its evaluation entails marginalising (i.e., integrating) out dependencies on model parameters. Technically, model evidence is accuracy minus complexity, where accuracy is the expected log likelihood of some data and complexity is the divergence between posterior and prior densities over model parameters.
**Variational bound** (a.k.a., **variational free energy**): known as an evidence lower bound (ELBO) in machine learning because it is always less than the logarithm of **model evidence**. In brief, variational free energy converts an intractable marginalisation problem—faced by sampling procedures—into a tractable optimisation problem. This optimisation furnishes the posterior density over model parameters and ensures the ELBO approximates the log evidence for a model: https://en.wikipedia.org/wiki/Variational_Bayesian_methods. Crucially, the variational bound includes an explicit estimate of model complexity, in contrast to the Akaike and Bayesian information criteria ^22^.
**Bayesian model comparison**: a procedure to compare different models of the same data in terms of **model evidence**. The marginal likelihood ratio of two models is known as a **Bayes factor**: https://en.wikipedia.org/wiki/Bayes_factor.
**Agent-based simulation models**: an alternative to equation-based models, usually used to simulate scenarios that are richer than models based upon population dynamics. Agent-based models simulate lots of individuals to create a sample distribution over outcomes. Evaluating the **marginal likelihood** from the ensuing sample distributions is extremely difficult, even with state-of-the-art estimators such as the harmonic mean: see https://radfordneal.wordpress.com/2008/08/17/the-harmonic-mean-of-the-likelihood-worst-monte-carlo-method-ever/.

## Acknowledgements

The Wellcome Centre for Human Neuroimaging is supported by core funding from the Wellcome Trust [203147/Z/16/Z].

## Author contributions

All authors contributed equally to the writing and revision of this work. The figures were prepared by KF.

## Author declarations

The authors declare no conflicts of interest. The authors are panellists on the Independent SAGE.

## Footnotes

i Government Publications: Research and analysis: COVID-19: Preparing for a challenging winter 2020/21, 7 July 2020 (Paper prepared by the Academy of Medical Sciences) https://acmedsci.ac.uk/file-download/51353957

ii https://www.gov.uk/government/publications/national-covid-19-surveillance-reports/sero-surveillance-of-covid-19

iii https://www.theguardian.com/world/2020/jun/07/immunological-dark-matter-does-it-exist-coronavirus-population-immunity

iv https://www.ons.gov.uk/peoplepopulationandcommunity/healthandsocialcare/conditionsanddiseases/bulletins/coronaviruscovid19infectionsurveypilot/18june2020#antibody-data

v https://www.gov.uk/government/publications/national-covid-19-surveillance-reports/sero-surveillance-of-covid-19

a https://www.fil.ion.ucl.ac.uk/spm/covid-19/

b Please see **DEM_COVID_I.m** for a Matlab demonstration that can be read as pseudocode (see software note).

c https://en.wikipedia.org/wiki/COVID-19_pandemic_on_Charles_de_Gaulle

